# Myosin Post-translational Modifications Associated with Critical Illness Myopathy

**DOI:** 10.1101/2025.08.18.25333907

**Authors:** Fernando Ribeiro, Bruno Di Geronimo, Nicola Cacciani, Anna Widgren, Yvette Hedström, Anselmo S. Moriscot, Peter M. Kasson, Shina C. L. Kamerlin, Jonas Bergquist, Lars Larsson

## Abstract

**Background:** Critical Illness Myopathy (CIM) is a devastating consequence of modern critical care, causing dramatic loss of muscle mass and function in intensive care unit (ICU) patients. However, the loss in function by far exceeds the loss in muscle mass and myosin content, but the molecular mechanisms underlying the loss of force and myosin-expressing non-force (NF) generating fibers remain elusive.

**Objectives:** To explore the mechanisms underlying the compromised myosin function in ICU patients exposed to 12-day mechanical ventilation and immobilization.

**Methods:** Mass spectrometry-based proteomics and molecular dynamics simulations were used to explore the pathophysiology underlying the compromised muscle fiber function previously reported at the single muscle fiber level in six ICU patients on 12^th^ day compared with the 1^st^ day of mechanical ventilation and immobilization.

**Results:** Previous measurements revealed single muscle fiber size and specific force to be decreased by ∼25% and 35% (p < 0.05), respectively, from the 1^st^ to the 12^th^ days. On the 12^th^-day, all fibers showed a decreased specific force and a subset of myosin-expressing fibers was identified exhibiting a complete loss of contractile function despite showing comparable fiber atrophy levels (∼30%, p < 0.05). Compromised force-generating capacity was linked to 27 post-translational myosin modifications, including oxidation, ubiquitination, acetylation, and methylation. Molecular dynamics simulations revealed an oxidative-induced rigidity of the myosin head, compromising the actin-binding and converter domains’ flexibility. Notably, the non-force generating fibers exhibited a unique proteomic signature linked with increased structural exposure and rigidity of the myosin motor domain.

**Conclusions:** In addition to muscle wasting and preferential myosin loss, abnormal myosin post-translational modifications contribute to the dramatic loss in muscle function in ICU patients with CIM, including the development of muscle fibers unable to generate contractile force.

## Introduction

Mechanical ventilation is a lifesaving intervention frequently used in modern critical care, but mechanical ventilation is also associated with complications related to lung injury, acquired myopathies, and cognitive dysfunction. The dramatic muscle wasting and compromised force-generating capacity of remaining contractile material negatively impact the rehabilitation of intensive care unit (ICU) patients and weaning off mechanical ventilation, resulting in prolonged ICU care with negative consequences for patient quality of life, mortality/morbidity, and health care costs (for refs. see (1)).

The loss of muscle mass and the preferential loss of the molecular motor protein myosin contribute to the decline in force generation capacity (maximum force normalized to muscle fiber cross-sectional area, or specific force) in limb muscles (1–3). In the diaphragm, the loss in specific force is not caused by a preferential loss of myosin, while myosin post-translational modifications negatively affect force-generation capacity (4, 5). The preferential myosin loss in limb muscles is a relatively late phenomenon in mechanically ventilated and immobilized ICU patients, i.e., the hallmark of the Critical Illness Myopathy (CIM), which is preceded by myosin post-translational modifications negatively affecting the specific force in limb muscles in a similar way as in the diaphragm (6).

In experimental studies, we have previously observed an increasing proportion of non-force generating diaphragm muscle fibers in rats mechanically ventilated from 6 hours to 14 days. In control diaphragm muscle fibers, all single muscle fibers generated normal specific force, while 7%, 22%, and 37% were non-force generating in animals immobilized and mechanically ventilated for 6 hours to 4 days, 5 to 8 days, and 9 to 14 days, respectively (4). In a more recent prospective clinical study involving mechanically ventilated neuro-ICU patients followed for 12 days with six repeated muscle biopsies were performed for transcriptome profiling. Additionally, contractile properties at the single muscle fiber level were measured in the first and final biopsy. In five of the 10 patients who survived 12 days of mechanical ventilation, between 9 and 21 % of the measured fibers did not generate any force upon maximum calcium activation despite significant myosin expression (2).

Thus, it is suggested that non-force generating fibers will have a significant impact on limb and respiratory muscle function in long-term immobilized and mechanically ventilated ICU patients, but underlying mechanisms remain elusive. We hypothesize that the non-force generating fibers have unique myosin modifications distinguishing them from control and force-generating fibers from long-term mechanically ventilated ICU patients. This hypothesis was supported by mass spectrometry-based proteomics and molecular dynamics simulation data.

## Materials & Methods

For details, see supplementary information.

### ICU patients

Six neuro-ICU patients (IDs #626, #629, #632, #639, #646 and #662) originated from a previous study (2) were included in this study. All patients exhibited a central nervous system injury and the anthropometric and medical history details were previously reported (2). Written consent was obtained from the patients’ close relatives and the study was approved by the ethics committee at the Karolinska Hospital (Dnr 2016/242-31/2).

### Muscle biopsies

Biopsies of the tibialis anterior (TA) muscle of patients were obtained on the 1^st^ (D1) and 12^th^ (D12) day of ICU hospitalization using the percutaneous conchotome method (2).

### Single muscle fiber size and specific force

Muscle bundles were dissected from the TA muscle and permeabilized for single fiber size and contractility measurements. Skinned single muscle fibers were carefully isolated and attached to force transducer connectors in the setup apparatus. Fiber’s absolute force (calculated as the difference between maximal isometric force and resting tension), cross-sectional area (CSA), and specific force (absolute force normalized to CSA) were determined.

### Mapping myosin PTMs with LC-MS/MS-based proteomics

Twenty-six muscle fibers isolated from ICU patients’ biopsies, distributed as follows: ICU_D1 (*n* = 4), ICU_D12 force-generating (*n* = 11), and ICU_D12 non-force-generating (*n* = 11), were used for liquid chromatography-mass spectrometry (LC-MS/MS) proteomics. Myosin protein content was separated on a 12% SDS-PAGE, and Coomassie-stained bands corresponding to myosin heavy chain (∼223 kDa) were excised, in-gel protein digested, and the peptides analyzed by LC-MS/MS for identification of PTMs.

### Molecular dynamics (MD) simulations

Human Myosin-7 (UniProt ID:P12883; PDB:4DB1) (7) constituted by two MYH7 chains (A and B) was selected as the model for our computational analysis. This high-resolution X-ray structure was modeled to reproduce the derived systems ‘’ICU_D1’’, ‘’ICU_D12’’ and ‘’ICU_D12_NF’’ by adding PTMs loop reconstruction, and ATP molecules, keeping the crystallographic water molecules (Figures E1-E2). The specific PTMs for each system were as follows: ICU_D1 included oxidized positions N589, D752, and H753; ICU_D12 featured oxidations at K86, D89, H97, Y162, Y164, H491, F494, N589, D752, and H753, as well as methylation at K757; and ICU_D12_NF included oxidation at H97. MD simulations were carried out using AMBER24 and AmberTools (7) using the ff19SB (8) and GAFF2 force fields (9). The three systems were initially energy-minimized *in vacuo* (10) and subsequently solvated in a truncated octahedral box using TIP3P (11) water molecules and adding counterions until charge neutrality. Three replicas of 500 ns conventional MD simulations were then performed for each system. Trajectory analysis was carried out using the CPPTRAJ module and Matplotlib (12).

### Statistical analysis

Data analysis was performed using Excel, Prism and R. The Shapiro-Wilk test was used to assess data normality. One-way analysis of variance (ANOVA) followed by Tukey’s post hoc tests, or Kruskal-Wallis’s followed by Dunn’s post hoc tests, was used for multiple comparisons between groups. Values are presented as means ± standard deviations unless stated otherwise. Statistical significance was accepted as *p* < 0.05.

## Results

It is hypothesized that differential regulatory control of myosin protein structure by distinct PTMs represents a molecular mechanism underlying the modulation of skeletal muscle weakness severity in ICU patients. To test this hypothesis, TA muscle biopsies were obtained from six neuro-ICU patients followed longitudinally for 12 days during immobilization and controlled mechanical ventilation previously described in detail (2). Single muscle fiber size, maximum force, and specific force (maximum force normalized to fiber CSA) previously characterized on the 1^st^ and 12^th^ day of immobilization and mechanical ventilation were used for proteomic analysis. LC-MS/MS-based proteomics was employed to examine myosin PTMs, followed by MD simulations for modeling the impact of PTMs on myosin protein structure and function in response to long-term ICU exposure (Figure 1A).

**Figure 1.**
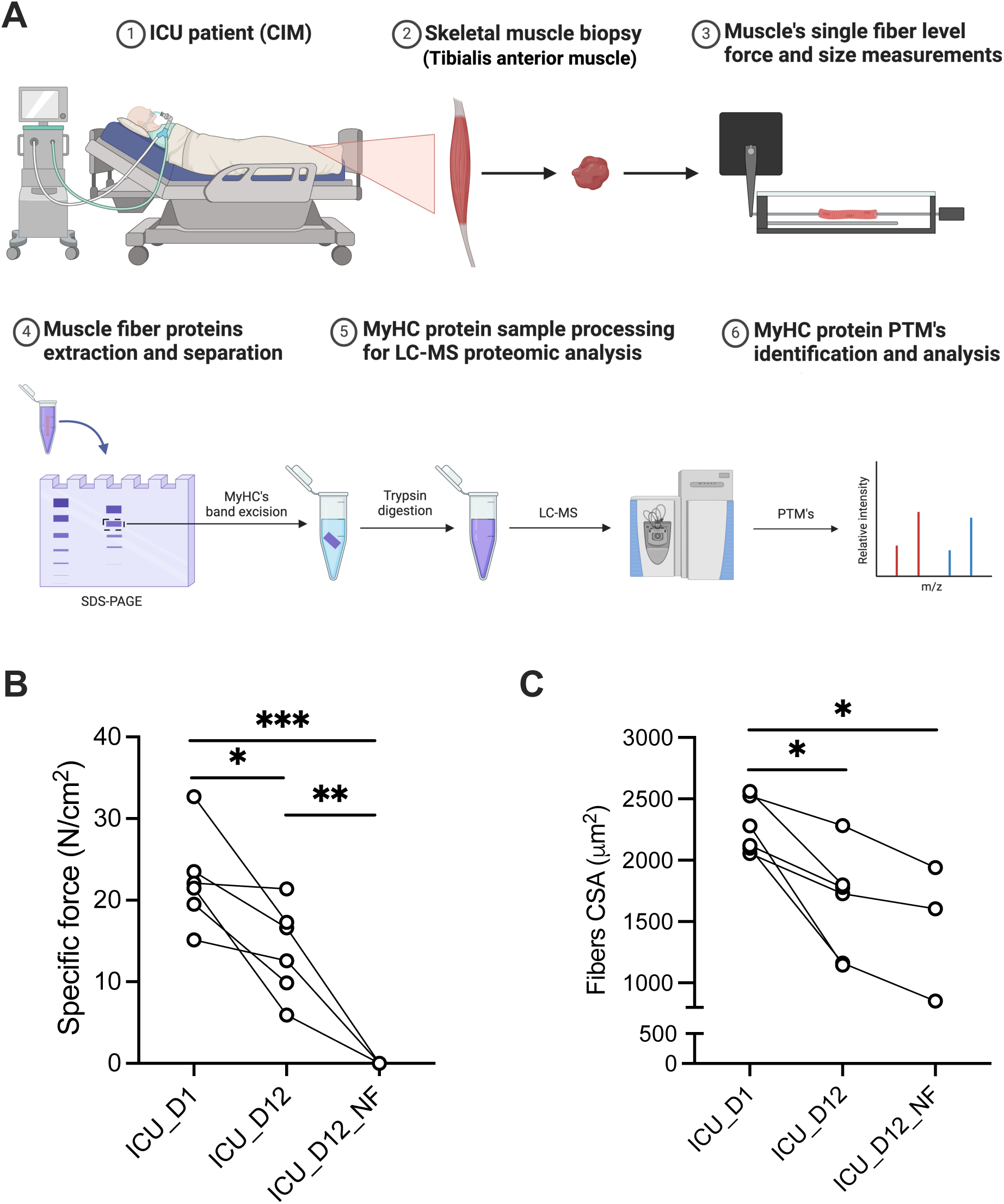
Investigating the role of PTMs on myosin as a regulatory molecular mechanism of the skeletal muscle weakness severity during critical illness myopathy. A) Schematic illustration of the experimental design employed in this study to investigate the impact of PTMs on myosin protein structure and function in muscle weakness during critical illness myopathy. B) ICU patient’s TA muscle specific force (i.e., the absolute force normalized to the fiber’s CSA), and fiber size (where each point represents the mean values, defined as the average specific force and fiber CSA measurements of 10-20 fibers, per subject, *n* = 3-5). The data presented for ICU_D1 and ICU_D12_F groups’ muscle fiber-specific force and size originate from previously published work (2), used for comparison with ICU_D12_NF group. Data are presented as mean ± SD. \**p* < 0.05; \*\**p* < 0.001; and \*\*\**p* < 0.0001 denote statistically significant differences among groups according to One-way ANOVA followed by Tukey’s post hoc test analysis. Graphical illustration was created with Biorender (biorender.com).

### ICU-induced muscle weakness and wasting

To investigate the negative impact of ICU treatment on skeletal muscle structure and function, we used the TA muscle fiber, whose contractile function and size were previously characterized at the single muscle fiber level (2). Significant decline in muscle fiber size and specific force was observed at the 12^th^ compared with the 1^st^, as previously reported (2) (see Figure 1B-C). Strikingly, in three out of six ICU patients, we identified a subpopulation of muscle fibers with a complete loss of force-generating capacity upon maximal calcium activation, despite showing comparable muscle fiber atrophy levels as in 12-day force-generating fibers (Figure 1C), and protein coverage of myosin motor protein (Figure 2A). These muscle fibers from individuals exposed to 12 days of mechanical ventilation and immobilization were categorized into 12-day force-generating (‘’ICU_D12’’) and 12-day non-force-generating (‘’ICU_D12_NF’’) muscle fibers.

**Figure 2.**
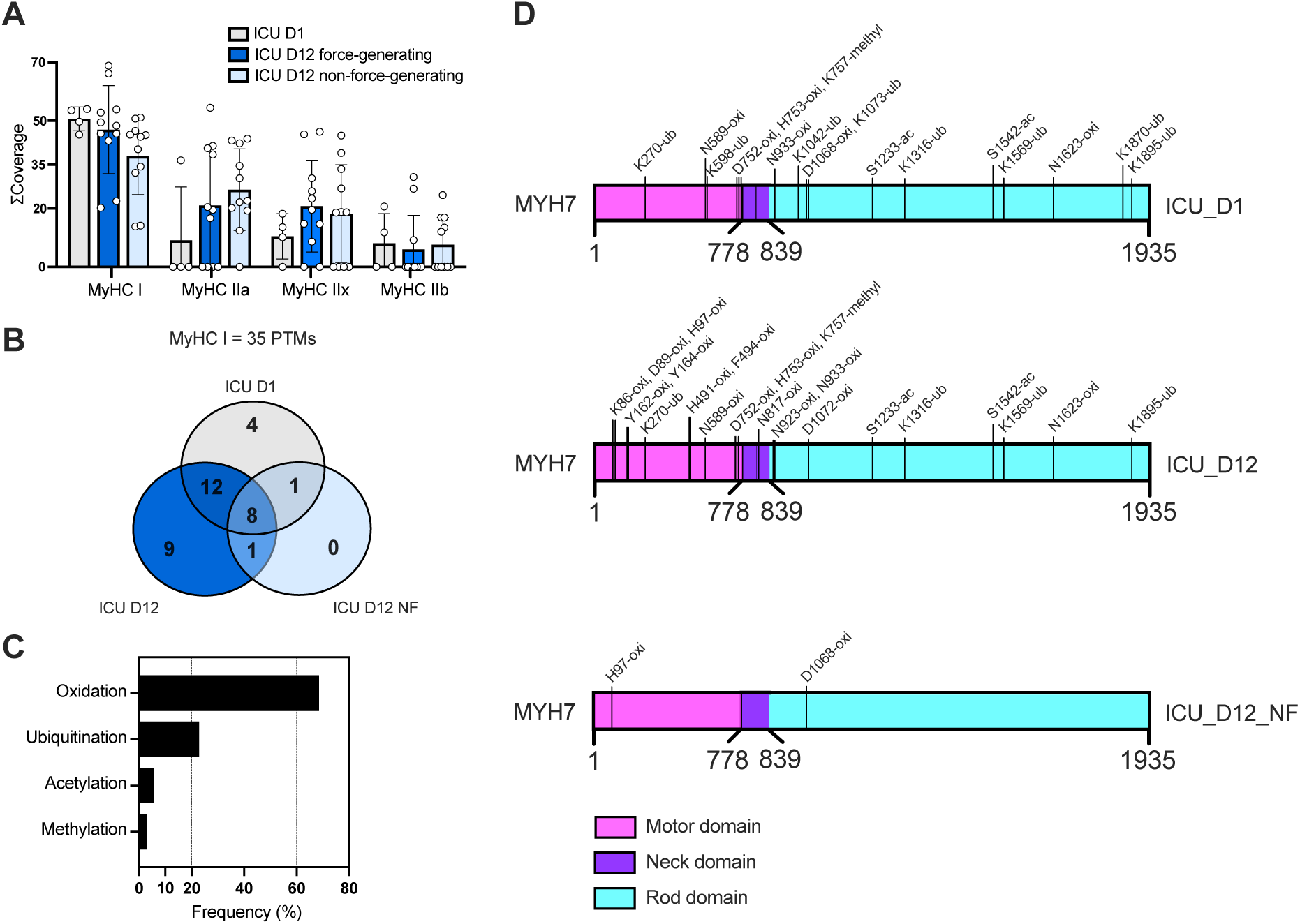
Post-translational modifications found in the tibialis anterior slow-twitch fibers of critically ill patients after 12 days of ICU hospitalization. A) Total coverage of myosin heavy chain protein isoforms isolated from the tibialis anterior muscle biopsies subjected to LC-MS/MS-based proteomic analysis. B) Venn diagram representing the amount of post-translational modifications identified and their distribution in each group. C) Relative frequency of distribution of post-translational modification per class found in the predominant MyHC type I isoform. D) MYH7 2D model representation depicting the localization of post-translational modifications present in the patient’s muscle fibers following one to twelve days of ICU hospitalization. Data are presented as mean ± SD, *n* = 4-11.

### Identification of myosin PTMs using LC-MS/MS-based proteomics

To investigate the mechanisms underlying the partial or total decline in specific force noted in fibers following 12 days of immobilization and mechanical ventilation, myosin protein PTMs were examined using LC-MS/MS-based proteomics. The presence and absence of myosin PTMs were analyzed in TA muscle fibers from ICU_D1 and compared with the respective ICU_D12 force- and non-force-generating groups.

Twenty-six muscle fibers across all experimental groups were included in this analysis. In humans, the TA muscle is predominantly composed of slow-twitch fiber, with approximately 80% of the fibers expressing the slow βslow/type I myosin heavy chain (MyHC) isoform (13). In line with this, all single muscle TA fibers included in this study predominantly expressed the type I MyHC isoform, although small amounts of fast MyHC isoforms (types IIa, IIx, and IIb) proteins were also detectable by mass spectrometry analysis (Figure 2A). Thus, the type I MyHC isoform dominated at the single muscle fiber level in the ICU_D1 and ICU_D12 groups, and there were no significant differences in MyHC isoform composition in the fibers analyzed in the three groups. Therefore, our PTM analyses were focused on the predominant type I MyHC isoform (Figure E1A). Overall, 35 PTMs of interest were identified, 8 of which were common across conditions, while the other 27 PTMs were differentially regulated between the ICU_D12 and ICU_D1 groups (Table 1). These 27 modifications can be separated into four major classes: 1) Oxidation (“Oxi’’, 68.6%); 2) Ubiquitination (“Ub’’, 22.9%); 3) Acetylation (“Ac”, 5.7%); and 4) Methylation (“Methyl”, 2.8%) (Figure 2B-2C). Ten PTMs (Lys86-Oxi; Asp89-Oxi; His97-Oxid; Tyr162-Oxi; Tyr164-Oxi; His491-Oxi; Phe494-Oxi; Ans817-Oxi; Ans923-Oxi; and Asp1072-Oxi) were added into ICU_D12 fibers (Figure E1B), with His97-Oxid also found present in ICU_D12_NF group, compared with ICU_D1 control group. Furthermore, 17 PTMs were absent in ICU_D12 groups compared with ICU_D1, of which four PTMs (Lys598-Ub; Lys1042-Ub; Lys1073-Ub; and Lys1870-Ub) that were absent in both ICU_D12, force- and non-force generating fibers, while one modification (Asp1068-Oxi) was found lacking only in the ICU_D12 group, compared with controls (Table 1). Strikingly, a total of 12 PTMs (Lys270-Ub; Asn589-Oxi; Asp752-Oxi; His753-Oxi; Lys757-Methyl; Asn933-Oxi; Ser1233-Ac; Lys1316-Ub; Ser1542-Ac; Lys1569-Ub; Asn1623-Oxi; and Lys1895-Ub) were exclusively absent in the ICU_D12_NF fibers group (Table 1).

**Table 1.**
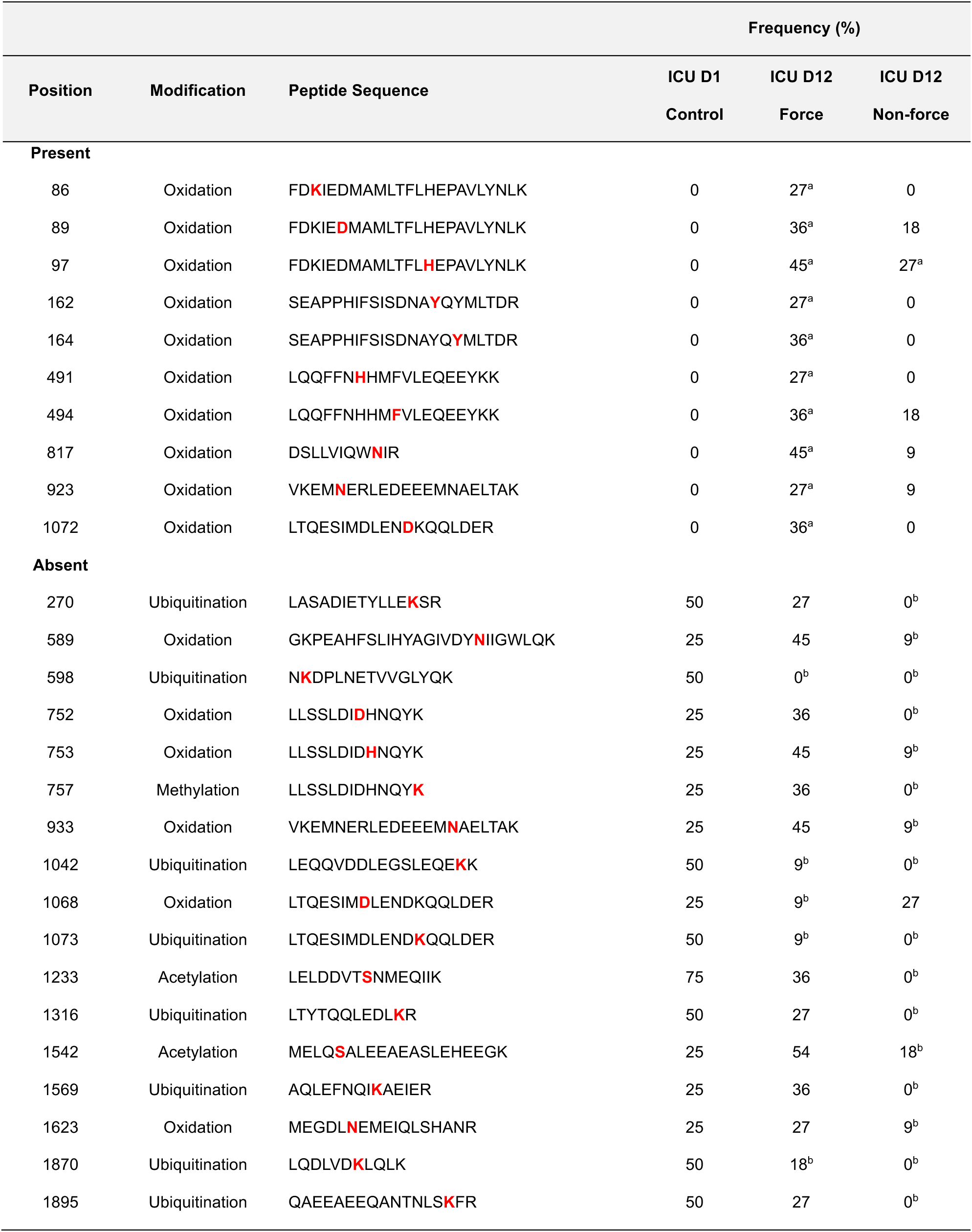
Identified post-translational modifications in MyHC type I protein. ^a^Indicates post-translational modifications with increased frequency compared with the control group; ^b^Indicates post-translational modifications with decreased frequency compared with control. The respective group’s sample sizes are ICU_D1 control (*n* = 4), ICU_D12 force-generating and ICU_D12 no force-generating fibers (*n* = 11).

Noteworthy, among the 10 novel PTMs found in ICU_D12 fibers, seven were located within the myosin protein motor domain, one in the neck, and the other two in the rod domain. Moreover, among the five missing modifications, one was located in the motor domain, while the remaining four were missing in the rod domain. On the other hand, ICU_D12_NF fibers showed only one newly added modification located within the myosin motor domain compared with control ICU_D1 (Figure 2D).

### Myosin molecular dynamics simulations

Analysis of trajectories from the conventional MD simulations showed a clear difference in behavior between the three simulated systems. Specifically, ICU_D1, displayed notable flexibility in the regions defined as the converter domain (amino acid index positions 655-677) and the actin-binding domain (amino acid index positions 757-771), being the latter one of the most flexible regions. Conversely, the main core of the myosin remained static (Figure 3). Notably, ICU_D12 and ICU_D12_NF exhibited reduced flexibility in both regions compared to ICU_D1 (Figure 3), and this increased rigidity was particularly pronounced in chain A (Figures E3-E4). A detailed analysis of position His97, located at the N-terminal region of helix-6, was carried out, showing a different behavior in the three groups. In ICU_D1, the non-oxidized histidine remained stable by keeping the two hydrogen bonds within the backbone of Ala100 and Gln78. In ICU_D12, His97 is oxidized to oxo-histidine, and despite the loss of the hydrogen bond with Gln78, the modified amino acids remained in the same position. However, in ICU_D12_NF fibers, this oxo-histidine was not stable and tended to be exposed to the solvent (Figure E5). In this system, the oxo-histidine was more flexible and moved apart from the position and tended to interact by pi-cation interaction with Arg706 (Figure E6). Solvent Accessible Surface Area (SASA) analysis allowed us to monitor this exposure, and the ICU_D12_NF group exhibited a more solvent-exposed His97 (Figure 4).

**Figure 3.**
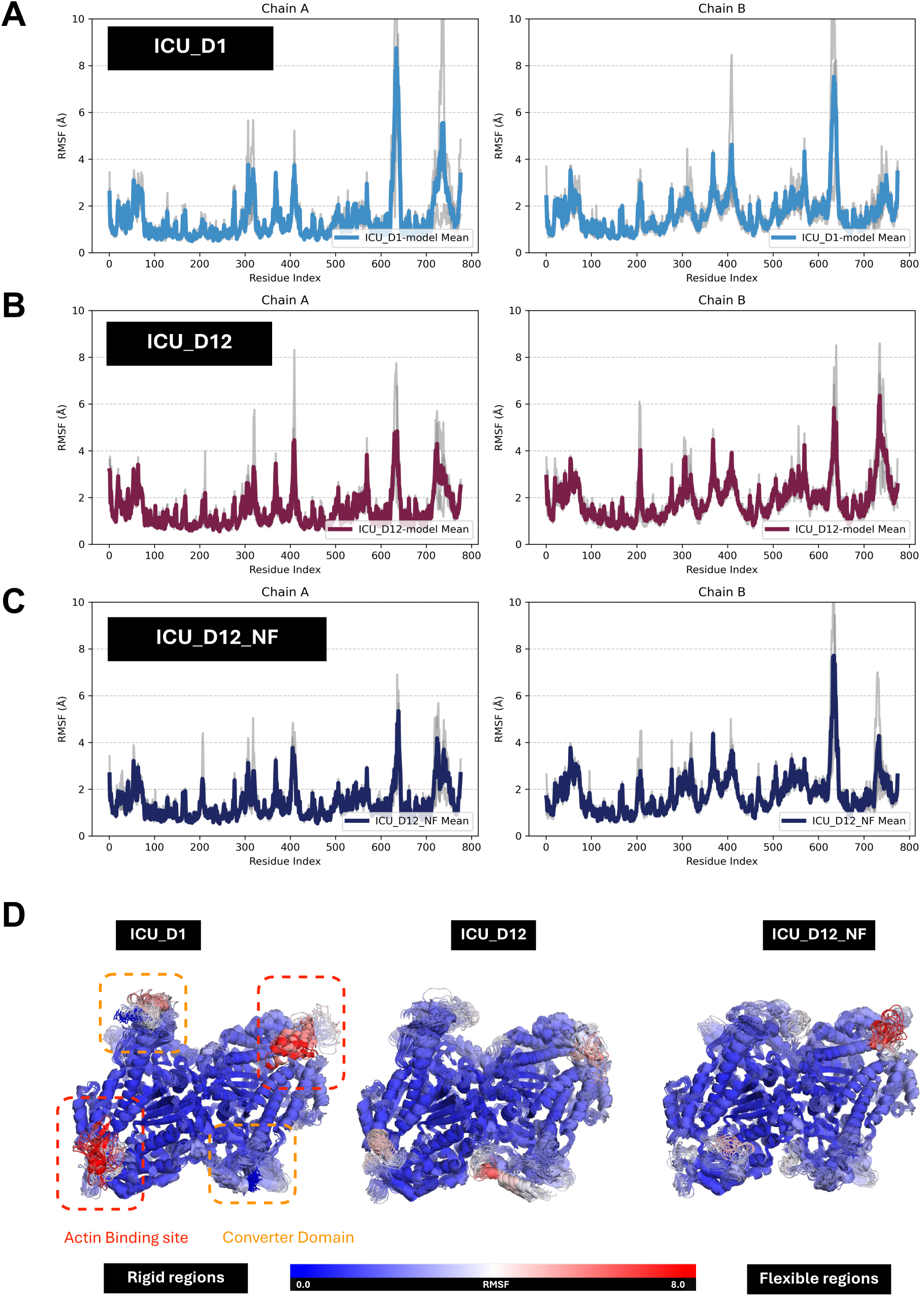
Prolonged mechanical ventilation and immobilization promote oxidative modifications and increased rigidity of myosin protein head. Residue RMSF values for each simulated replica (gray lines) and the corresponding mean RMSF (colored lines) for systems A) ICU_D1, B) ICU_D12, and (C) ICU_D12_NF, shown separately for chains A and B. D) Molecular dynamics RSMF analysis confirms elevated flexibility of actin-binding and converter domains in the ICU_D1 control group, while after 12 days of ICU treatment (mechanical ventilation and immobilization) these domains critical domains required for myosin contractile function show decreased flexibility (i.e. rigidity).

**Figure 4.**
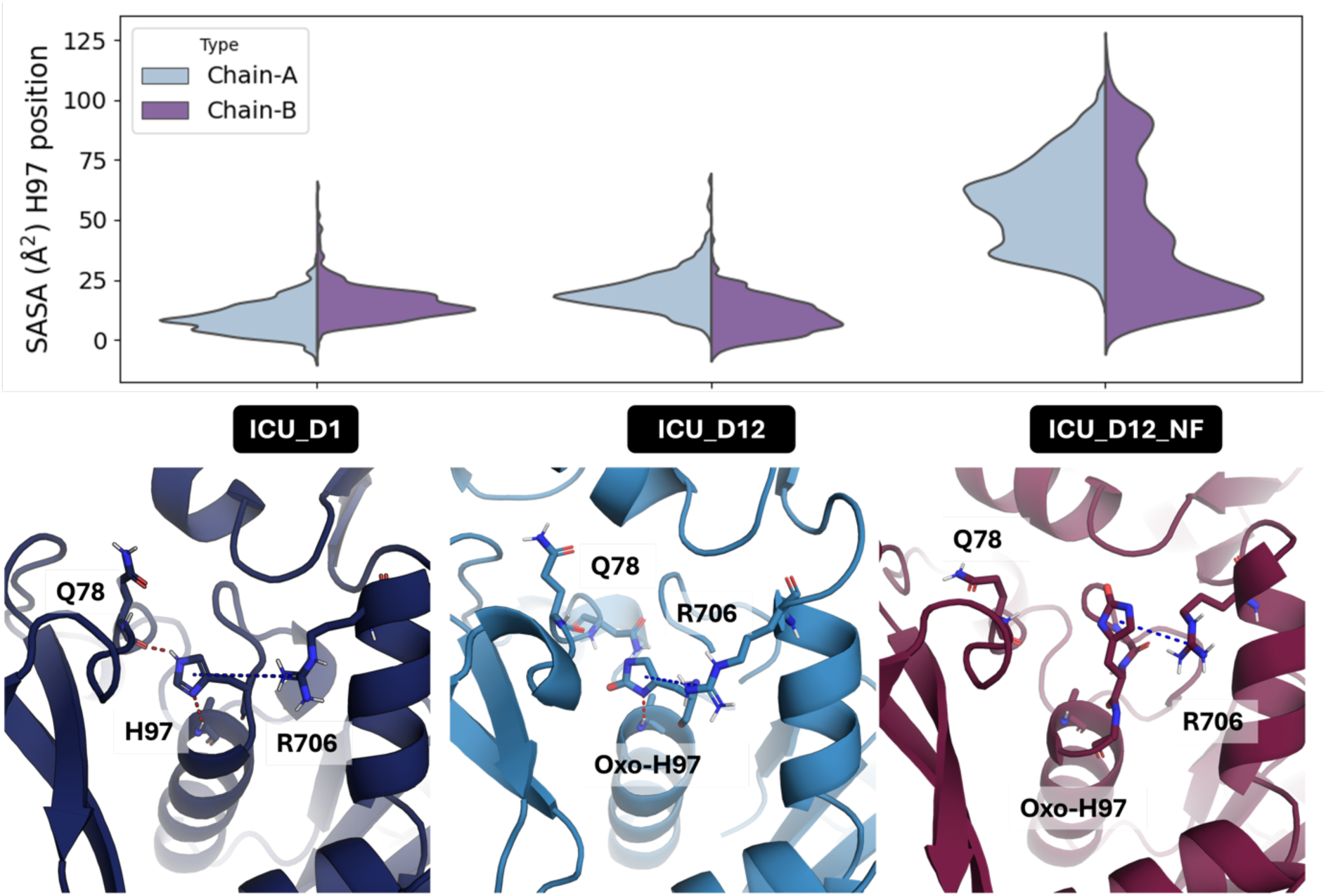
Non-force generating fibers show a unique structural signature with higher exposure and instability of myosin motor domain induced by H97 oxidation. The solvent-accessible surface area (SASA) analysis suggests that H97 oxo-histidine is not stable and tends to be exposed to the solvent in non-force generating fibers. In this system, the H97 residue is more flexible, moving apart from the position, and tends to interact by pi-cation interaction with the Arg706, which may negatively impact the movement of the myosin converter domain during the powerstroke state of muscle contraction.

## Discussion

Critically ill patients undergoing prolonged mechanical ventilation and immobilization rapidly develop skeletal muscle wasting and weakness (14–16). Muscle biopsies from ICU patients reveal significant ultrastructural changes, including impaired thick filament organization and preferential myosin loss (2, 17–20). However, the molecular mechanisms underlying myosin contractile dysfunction and loss remain unknown. Herein, we examined the effects of prolonged ICU treatment (i.e. mechanical ventilation and immobilization) on myosin post-translational modifications and their role in muscle weakness. Limb muscle biopsies from a previous study (2) collected from neuro-ICU patients were reanalyzed with proteomic and molecular dynamics simulation analyses to assess myosin post-translational modifications and their impact on protein structure and function in relation to muscle fiber size and specific force on the 12^th^ compared with the 1^st^ day of immobilization and mechanical ventilation. Twelve days of mechanical ventilation and immobilization had a strong negative effect on muscle fiber size and specific force (2) coupled to post-translational myosin modifications. Abnormal post-translational modifications (e.g., oxidation, ubiquitination, acetylation, and methylation) were associated with myosin protein rigidity and contractile dysfunction. Additionally, a subset of fibers exhibited complete loss of contractility, showing a unique proteomic signature characterized by dramatically fewer post-translational modifications, predicted to destabilize myosin motor domain structure and function.

Elevated oxidative stress is known to impair skeletal muscle contractile function (21). *In vitro* and *in vivo* studies show that oxidative modifications induce sarcomere protein damage due to altered protein ultrastructure, protein-protein binding interaction, and calcium sensitivity (22). In parallel with the decline in specific force after 12 days of immobilization and mechanical ventilation, multiple myosin post-translational modifications were observed, with oxidation being the most prevalent one, accounting for over 60% of all identified modifications. Prolonged mechanical ventilation and immobilization are associated with inflammation and mitochondrial dysfunction, both known to enhance reactive oxygen species (ROS) production in critically ill patients (23). Our previous experimental studies confirm that long-term mechanical ventilation and immobilization ROS-induced modifications (e.g. oxidation, carbonylation) impair efficiency, motility speed, and force-generation capacity of myosin at the motor protein level (4, 5, 24). Thus, mechanisms underlying muscle dysfunction in ICU patients with CIM are multifactorial and not only related to muscle fiber atrophy, altered membrane excitability and excitation-contraction coupling, and preferential myosin loss but also post-translational myosin modifications compromising function as well as a subset of non-force generating muscle fibers.

ICU patients’ myofibers exhibit increased oxidative stress-induced myosin post-translational modifications (6, 18), but how these modifications disturb myosin structure and contractile function remains unclear. Observations from the current study demonstrate increased rigidity of the myosin motor domain in response to oxidative modifications after 12 days of ICU treatment, affecting myosin actin-binding and converter domains. Structural analysis revealed proximity between oxidized residues and methionine residues (Figure E7), suggesting methionine oxidation may trigger the oxidative process, as previously described (25, 26). Recent data suggest an increased fraction of myosin protein trapped in the super-relaxed state (i.e., blocked myosin-actin binding interaction) (27). However, this phenomenon was uniquely observed in the diaphragm and thus unlikely to explain the limb muscle weakness observed herein. Conversely, myosin oxidative modifications have been noticed in both respiratory and limb muscles following prolonged mechanical ventilation (4, 6, 18). Thus, oxidized sites located close to each other in the converter domain are suggested to affect the myosin lever arm movement and decrease myosin head flexibility, ultimately impairing myosin function in ICU patients.

Prolonged mechanical ventilation leads to the emergence of non-force generating fibers, contributing to muscle weakness severity in critically ill patients. We previously reported in a time-course experimental study a progressive increase of this subset of fibers, from 7% to 37%, in the diaphragm after 6 hours to 14 days of mechanical ventilation and immobilization (4) and confirmed in a prospective clinical study showing 9–21% non-force generating limb muscle fibers in 5 of 10 neuro-ICU patients exposed to 10-12 days mechanical ventilation and immobilization (2). In this study, non-force generating fibers had similar fiber CSA and myosin content as in force generating ICU D12 fibers. However, these dysfunctional fibers uniquely exhibited increased solvent-accessible surface area at oxidized histidine 97, suggesting higher solvent exposure, pointing to a structural rearrangement upon oxidation. While increased solvent exposure does not inherently imply reduced structural stability of the myosin head, detailed analysis reveals a newly reinforced π–cation interaction between His97 and Arg706, observed exclusively in the non-force-generating fiber.

Interestingly, this particular histidine oxidation was also detected in fast-twitch fibers (type IIx) in the diaphragm of rats exposed to 5 days of mechanical ventilation (28). This particular histidine (i.e., H98) was also found oxidized in the detectable fraction of myosin type IIx protein from non-force generating fibers (Table E1, Figure E8). One may envision that oxidized histidine residues may serve as electrophiles and readily react with nearby nucleophiles to form covalent adducts, leading to the complete loss of the lever arm movement and explaining the complete loss of myosin contractile function observed in ICU_D12_NF fibers. This aligns with previous evidence demonstrating that exacerbated and cumulative oxidative stress may trigger the transition of cross-bridges from the force-generating to non-force generating state (29). Hence, we propose that non-force generating fibers represent a subpopulation of fibers in an advanced degenerative state resulting from the cumulative oxidative damage induced by prolonged mechanical ventilation and immobilization.

Although oxidation was predominant, other identified post-translational modifications may also affect myosin in ICU patients. In this context, protein ubiquitination plays a crucial role in several biological processes (e.g., muscle proteostasis, mitophagy, and regeneration) (30, 31). We found eight ubiquitinated sites in the myosin molecule of ICU_D1 controls, while after 12 days, four ubiquitinated sites were lacking in ICU_D12 fibers, mostly in the rod domain. Remarkably, all identified ubiquitinated sites were found missing in non-force generating fibers. To the best of our knowledge, this is the first report of altered ubiquitylation of human myosin muscle protein after long-term ICU treatment. Previous studies have shown increased activation of some ubiquitin-proteasome pathway markers in critically ill patients undergoing thoracic surgery, thought to contribute to muscle atrophy in mechanically ventilated patients (32). Concerning this, however, our proteomic supports that these contractile dysfunctional fibers expressed the full-length myosin protein (i.e. 223 kDa), suggesting the protein peptide sequence integrity in these biopsies. In our view, these altered ubiquitinated sites are most likely mono-ubiquitinations, which are highly dynamic and commonly involved with the regulation of enzymatic activity rather than protein degradation (33, 34). Thus, we envision that these lost ubiquitinations could negatively impact on myosin structure and function in long-term mechanically ventilated and immobilized ICU patients.

Acetylation represents another class of post-translational modifications that regulate muscle contraction in health and disease (35). We identified two lacking acetylations (Ser1233-Ac and Ser1542-Ac) in the myosin rod domain of myosin in non-force generating fibers. Similar acetylation modifications in the myosin rod domain have been reported in previous experimental and clinical ICU studies (18, 28). Decreased acetylation of contractile proteins has also been reported as a hallmark in disuse-induced muscle wasting and weakness (36). In line with this, histone deacetylases (HDACs), particularly HDAC4, have been demonstrated to target the myosin-heavy chain and promote muscle dysfunction (37). Missing acetylation sites in the myosin rod domain can disrupt thick filament stability, myosin head positioning and the ATPase turnover, contributing to muscle dysfunction in diseases such as congenital myopathies and heart failure (38, 39). Therefore, the lost acetylations in the myosin rod domain may contribute to the impaired myofibers’ force generation capacity in critically ill patients.

Protein methylation also controls important biological processes (40). Here, we identified a missing methylation (C-term K757) from the myosin motor domain of non-force generating fibers. Previously, our group reported myosin methylation changes in both experimental and clinical ICU studies (6, 18). These modifications have been suggested to be associated with impaired myosin filament stability, assembly, and motility, similar to aging-related methylation myosin modifications in humans (41). Furthermore, we have previously shown that BGP-15 treatment protects the soleus muscle strength in parallel with mitigating the methylation of the myosin rod domain in rats subjected to prolonged mechanical ventilation and immobilization (6). However, the understanding of the impact of methylation on myosin structure and function remains incompletely understood. Overall, the altered methylation in the myosin head (e.g. K757-methylation) may contribute to the loss of myosin function in non-force-generating fibers.

Study limitations: 1) Our patient sample size (n = 6) and the number of muscle fibers (n = 26) analyzed are relatively small due to the challenge of obtaining intra-subject matched muscle biopsies in long-term ICU patients. However, to our knowledge, this is the first clinical study that provides mechanistic insights into how myosin post-translational modifications negatively affect skeletal muscle contractile function in long-term (12 days) mechanically ventilated ICU patients. Furthermore, all muscle samples were collected from neuro-ICU patients with relatively similar clinical conditions (i.e., central nervous system injury) without underlying chronic disease and intrasubject time-course comparisons (ICU day 12 vs day 1), minimizing the inherent variability between subjects’ responses to long-term ICU treatment. 2) Other myofibrillar proteins besides the molecular motor protein myosin may also be affected by post-translational modifications may also influence force-generation capacity. However, myosin-actin interactions represent the final functional contractile unit and represent a key factor in the motor system. 3) Our molecular dynamics simulations are based on the myosin crystal structure (PDB: 4DB1) that contains some unresolved structural loops, and computational reconstruction of these missing loops remains uncertain, relying on predicted conformations rather than structural data.

In conclusion, abnormal post-translational myosin modifications (e.g., oxidation, ubiquitination, acetylation and methylation) are associated with muscle weakness development in neuro-ICU patients. Our findings suggest that oxidative modifications increased myosin head rigidity, affecting the actin-binding and converter domains, impairing myosin function. Moreover, a subset of non-force generating fibers exhibited a unique proteomic signature linked to increased instability and rigidity of the myosin protein motor domain. Therefore, the development of non-force generating fibers likely exacerbates muscle weakness in critically ill patients. These findings may provide a rationale for the design of novel therapies targeting myosin structure and contractile function preservation in long-term mechanically ventilated ICU patients.

## Author’s contributions

LL, NC, YH organized and conducted the clinical study together with single muscle fiber contractile measurements, and prepared single muscle fiber myosin for proteomics analyses. FR, AW, ASM and JB conducted mass spectrometry analyses. BDG, PMK and SCLK conducted modelling experiments. FR, BDG and LL drafted the original manuscript. All authors were involved in reviewing and finalizing the manuscript.

## Data availability statement

All data produced in the present study are available upon reasonable request to the corresponding author.

