## Supplementary information for "Myosin Post-translational Modifications Associated with Critical Illness Myopathy"

### Material & Methods

#### ICU patients

Six neuro-ICU patients (IDs #626, #629, #632, #639, #646 and #662) were included in this study (four male and two female subjects ranging in age between 26 and 83 years old). Further details about the patient's anthropometric and medical history were previously reported (1). Written consent was obtained from patient's close relative and the study was approved by the ethics committee at the Karolinska Hospital (Dnr 2016/242-31/2).

#### Muscle biopsies

Muscle biopsies of the tibialis anterior (TA) muscle of patients were obtained on the 1st (D1) and 12th (D12) days of ICU hospitalization (mechanical ventilation and immobilization) using the percutaneous conchotome method. Then, the samples were used for functional and morphometric measurements (as described below), snap-frozen in propane chilled by liquid nitrogen, and stored at -140°C for further analysis.

#### Muscle bundles permeabilization

Bundles (approximately 50 fibers) were dissected from the TA muscle in a relaxing solution at 4°C and tied to glass capillaries, stretched about 110% of their resting slack length. Next, the bundles were chemically skinned by treatment for 24 hours at 4°C in a relaxing solution containing 50% (v/v) glycerol, and then stored at −20°C. Within 1 week following skinning, the bundles were cryoprotected by treatment with relaxing solutions containing increasing concentrations of sucrose, (0, 0.5, 1.0, 1.5, and 2.0 M) for 30 minutes each step. Then, the cryoprotected bundles were snap-frozen in propane chilled with liquid nitrogen, and stored at -140°C until further use.

#### Skinned single muscle fiber size and function analysis

On the day of fiber size and contractile measurements, muscle bundles were transferred from -140°C to 2M sucrose solution, and incubated in decreasing sucrose concentration solutions (1.5-0.5 M) for 30 min each step. Finally, the bundle was transferred to the skinning solution kept at -20°C. Then, skinned single muscle fibers were carefully isolated and attached to force transducer connectors in the setup apparatus (for details see (2)). The fiber absolute force (calculated as the difference between maximal isometric force and resting tension), cross-sectional area (CSA), and specific force (absolute force normalized to CSA) were determined.

#### Mapping myosin post-translational modifications with mass spectrometry proteomics

Twenty-six muscle fibers from three ICU patients distributed as: ICU D1 ($n$ = 4), ICU D12 force-generating ($n$ = 11), and ICU D12 non-force-generating ($n$ = 11) were used for liquid chromatography tandem mass spectrometry (LC-MS/MS) proteomics analysis of myosin PTMs. Total MyHC proteins were separated on a 12% SDS- PAGE and Coomassie-stained bands corresponding to MyHC (~223 kDa) were excised, subjected to in-gel protein digestion, and peptides analyzed by LC-MS/MS (as described below).

### LC-MS/MS

The samples were analyzed using a Q Exactive Plus Orbitrap mass spectrometer (Thermo Fisher Scientific, Bremen, Germany) equipped with a nano-electrospray ion source. The peptides were separated via reversed-phase LC using an EASY-nLC 1000 system (Thermo Fisher Scientific). A set-up of a pre-column and an analytical column was used. The pre-column was a 2-cm EASY-Column (ID 100 µm, 5 µm C18;

Thermo Fisher Scientific) and the analytical column was a 10-cm EASY-Column (ID 75 µm, 3 µm, C18; Thermo Fisher Scientific). Peptides were eluted with a 35 min linear gradient from 4% to 100% acetonitrile at 250 nL min−1. The mass spectrometer was operated in positive ion mode, acquiring a survey mass spectrum with resolving power 70,000 (full-width half maximum), m/z = 302–1750, using an automatic gain control target of 3 × 106. The 10 most intense ions were selected for higher-energy collisional dissociation fragmentation (25% normalized collision energy) and MS/MS spectra were generated with an automatic gain control target of 5 × 105 at a resolution of 17,500. The mass spectrometer operated in data-dependent mode.

**Identification of Myosin Post-translational Modifications**

The acquired RAW data files were analyzed with the Proteome Discoverer 1.4.0.288 (Thermo Fisher Scientific) software using the SEQUEST HT® (University of Washington) search engine against proteins from Rattus norvegicus in the UniProtKB/SwissProt database downloaded June, 2022. The search parameters included the following: maximum 10 ppm and 0.6 Da error tolerance for the survey scan and MS/MS analysis, respectively; enzyme specificity was trypsin; maximum of two missed cleavage sites allowed; cysteine carbamidomethylation was set as static modification; oxidation (M) and deamidation (N,Q) were set as variable modifications. To search for many post-translational modifications (PTMs), the processing was performed in four blocks with the following variable modifications: (1) carbonylation (E, I, K, L, Q, R, T, V); (2) oxidation (F, H, K, P), phosphorylation (S, T, Y); (3) oxidation (D, N, R, W, Y); (4) ubiquitination (K), methylation (C-terminal), acetylation (K, S), nitration (W, Y). The search results from these blocks were merged and validated using Percolator (embedded in Proteome Discoverer). The protein identifications were based on at least two matching peptides per protein.

### Molecular dynamics (MD) simulation

### Myosin Systems Preparation

ICU_D1, ICU_D12 and ICU_D12_NF were built based on crystal structure 4DB1 (Figure E1). This structure presents missing loops at positions 403-410, 626-644, and 715-742 that were added using Modeller (3). PTMs were added as next:

- ICU_D1: oxidized positions N589, D752 and H753
- ICU_D12: oxidized positions: K86, D89, H97, Y162, Y164, H491, F494, N589, D752 and H753; methylated: K757
- ICU_D12_NF: oxidized positions: H97

Oxidized modifications have been done according to Figure E2, based on the available experimental data and literature (4–9).

To get the corresponding parameters from these non-canonical amino acids, as well as ATP molecules, we calculated at the HF/6-31G(d) level of theory using restrained electrostatic potential (RESP) (10) writing with Antechamber (GAFF2) (11), based on gas-phase geometries optimized at the B3LYP/6-31G(d) level of theory using Gaussian (Gaussian 16). Once the systems were parametrized using tleap module from AmberTools (12). After that, protein canonical residues were parameterized using the AMBER ff19SB (13) using H++ (14) to get titratable residues protonation state and crystallographic water molecules were parametrized using TIP3P model (15). Corresponding topology files and initial coordinates were built, after which the systems underwent energy minimization in vacuo. The minimized structures were subsequently solvated in a truncated octahedral box of TIP3P waters (72.000-73.000), and counterions were added to achieve charge neutrality.

**Conventional MD simulations:**

Molecular dynamics (MD) simulations were performed using the AMBER24 software using similar protocol. The three systems previously parametrized underwent energy minimization to eliminate steric clashes with the solvent and ions added. Simulations proceeded through an initial heating phase from 100 K to 300 K under the NVT ensemble, applying positional harmonic constraints (40 kcal·mol$^{-1}$·Å$^{-2}$) on solute atoms. These constraints were progressively reduced to 10 kcal·mol$^{-1}$·Å$^{-2}$ over four subsequent equilibration stages at constant temperature (NVT, 300 K) and eventually removed for the final equilibration under the NPT ensemble at 300 K. Each system was then subjected to three independent MD simulations of 500 ns each, totaling 1.5 μs per system. A 12 Å cutoff was used for van der Waals interactions, while long-range electrostatics were computed using the Particle-Mesh Ewald (PME) method. Bonds involving hydrogen atoms were constrained using the SHAKE algorithm. MD trajectories were analyzed using AmberTools software (12) to determine structural and dynamical parameters, including atom-positional metrics such root-mean-square deviations (RMSD), as root-mean-square fluctuations (RMSF), Solvent-Accessible Surface Area and distances using CPPTRAJ module. RMSF, SASA, and distance measurements were calculated using the last 300 ns of the simulations. Figures were done with PyMOL (pymol.org) and plot and statistical analysis were carried out using Matplotlib (16). The computational dataset and analysis supporting this study are available at Zenodo (https://doi.org/10.5281/zenodo.15278114).

**Statistical analysis**

Data analysis was performed using Excel, Prism and R. Shapiro-Wilk test was used to assess data distribution. One-way analysis of variance (ANOVA) followed by

Tukey's post hoc tests or Kruskal-Wallis's followed by Dunn's post hoc tests was used for multiple comparisons of data with normal or non-normal distribution, respectively. Values are presented as mean ± standard deviation (SD) unless indicated differently. Statistical significance was accepted as $p < 0.05$.

### Results

#### Table Legend

**Table E1.** [a]Indicates post-translational modifications with increased frequency compared with the control group; [b]Indicates post-translational modifications with decreased frequency compared with control. The respective group's sample sizes are ICU_D1 control (*n* = 4), ICU_D12 force-generating (*n* = 11) and ICU_D12 no force-generating fibers (*n* = 11).

#### Figures Legend

**Figure E1.** (A) General structure of Myosin MYH7 (PDB ID: 4DB1). Chain A is represented in cartoon style, while Chain B is shown as a surface. The ATP molecule is shown in green sticks, and water molecules are also represented. (B) Detailed view of MYH7 Chain A highlighting the positions and identities of amino acids selected for oxidative PTM modifications.

**Figure E2.** Schematic 2D representation of the proposed oxidation states for each amino acid.

**Figure E3.** Backbone RMSD values for each simulated replica (gray lines) and the corresponding mean RMSD (colored lines) for systems ICU_D1, ICU_D12, and ICU_D12_NF, shown separately for chains A and B.

**Figure E4.** Comparison of ΔRMSF values for ICU_D1 vs ICU_D12, and ICU_D1 vs ICU_D12_NF, shown separately for chains A and B.

**Figure E5.** Representative conformations of ICU_D1, ICU_D12, and ICU_D12_NF extracted from MD simulations, highlighting the distinct conformational states and interactions involving Histidine 97.

**Figure E6.** KDE plots showing the relationship between Histidine 97 RMSD and the distance between Histidine 97 and Arginine 706 from MD simulations of ICU_D1, ICU_D12, and ICU_D12_NF. The region enclosed by red dashed lines highlights the stable π–π interaction observed specifically in the ICU_D12_NF system.

**Figure E7.** General structure of Myosin MYH7 Chain A (PDB ID: 4DB1) represented in cartoon style, with selected amino acids highlighted in stick representation. Panels illustrate the spatial proximity between oxidizable residues and nearby methionines. (A) Lysine 86 and Methionine 113; (B) Aspartic acid 89 and Methionine 92; (C) Histidine 97 and Methionine 77; (D) Tyrosines 162 and 164 with Methionine 165; (E) Histidine 491 and Phenylalanine 494 with Methionine 515; (F) Aspartic acid 752 and Histidine 753 with Methionine 776.

**Figure E8.** Schematic 2D representation of identified post-translational modification and their localization in the structure of type IIx myosin protein for ICU_D1, ICU_D12, and ICU_D12_NF groups.

### Supplementary Tables

**Table E1**. Identified post-translational modifications in MyHC type IIx protein.

| | | | Frequency (%) | | |
|---|---|---|---|---|---|
| **Position** | **Modification** | **Peptide Sequence** | **ICU D1 Control** | **ICU D12 Force** | **ICU D12 Non-force** |
| **Present** | | | | | |
| 98 | Oxidation | YDKIEDMAMMTHL**H**EPAVLYNLK | 0 | 0 | 28[a] |
| 494 | Oxidation | LQQFFN**H**HMFVLEQEEYKK | 0 | 33[a] | 0 |
| 497 | Oxidation | LQQFFNHHM**F**VLEQEEYKK | 0 | 44[a] | 28[a] |
| **Absent** | | | | | |
| 154 | Oxidation | RQEAPP**H**IFSISDNAYQFMLTDR | 33 | 22[b] | 0[b] |
| 165 | Oxidation | RQEAPPHIFSISDNAYQ**F**MLTDR | 33 | 22[b] | 14[b] |
| 273 | Ubiquitination | LASADIETYLLE**K**SR | 66 | 22[b] | 0[b] |
| 601 | Ubiquitination | N**K**DPLNETVVGLYQK | 66 | 0[b] | 0[b] |
| 1046 | Ubiquitination | LEQQVDDLEGSLEQE**K**K | 66 | 11[b] | 0[b] |
| 1200 | Oxidation | **H**ADSVAELGEQIDNLQR | 33 | 0[b] | 0[b] |
| 1541 | Oxidation | QVEQE**K**SELQAALEEAEASLEHEEGK | 66 | 22[b] | 42 |
| 1635 | Oxidation | MEGDLNEMEIQLN**H**ANR | 33 | 22[b] | 0[b] |

### Supplementary Figures

**Figure E1.**

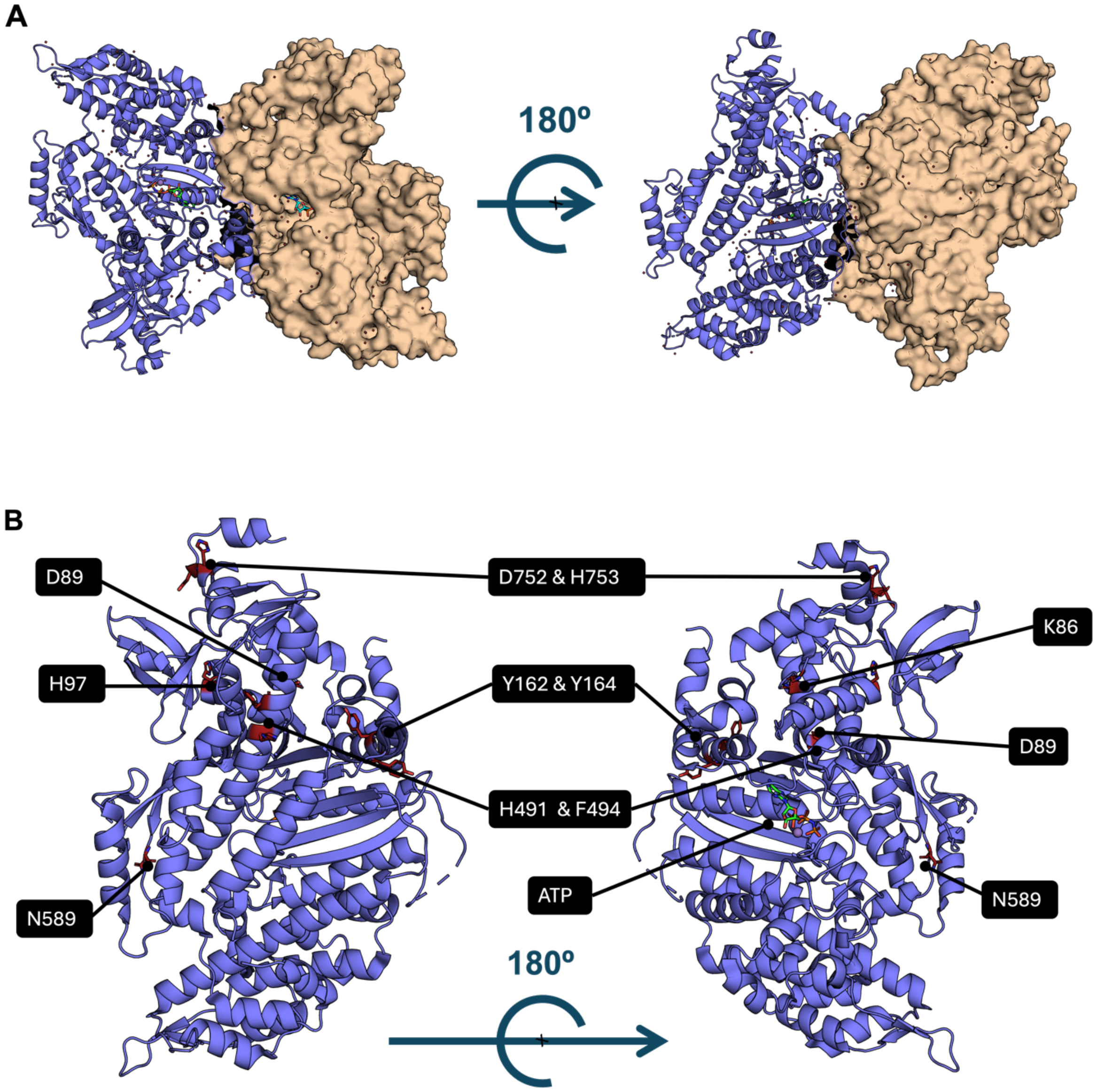

**Figure E2.**

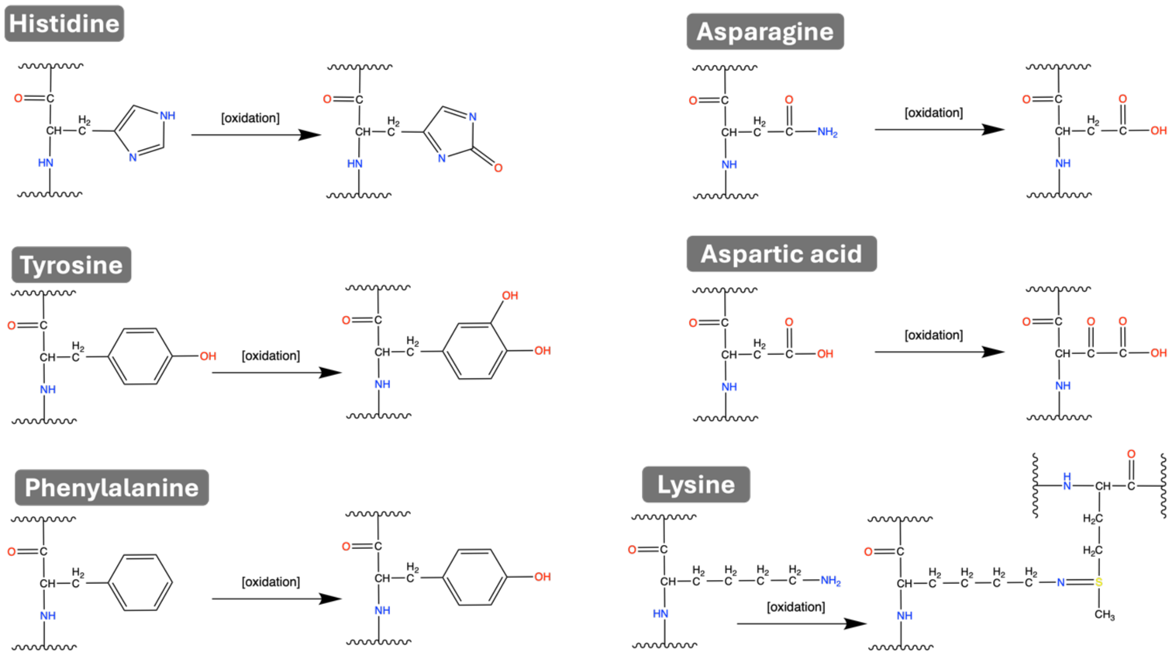
Histidine
[oxidation]
Asparagine
[oxidation]
Tyrosine
[oxidation]
Aspartic acid
[oxidation]
Phenylalanine
[oxidation]
Lysine
[oxidation]

### Figure E3.

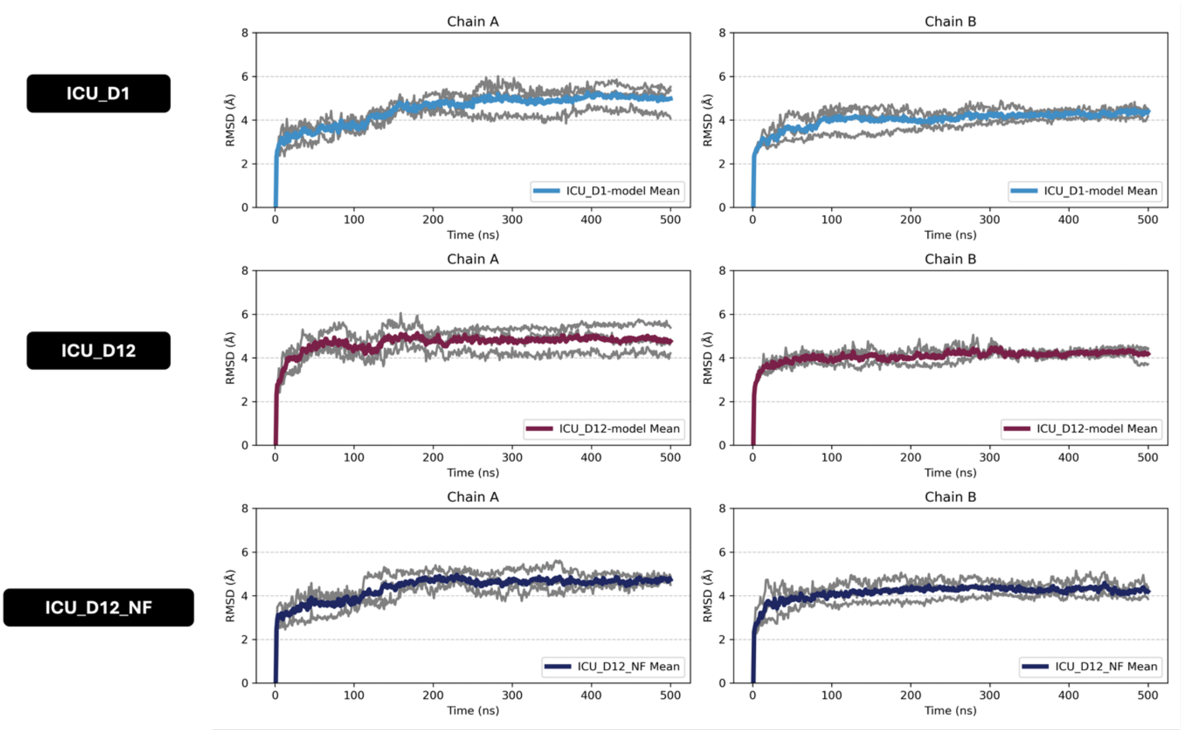
ICU_D1
ICU_D12
ICU_D12_NF
Chain A
Chain B
RMSD (Å)
Time (ns)
ICU_D1-model Mean
ICU_D12-model Mean
ICU_D12_NF Mean

**Figure E4.**

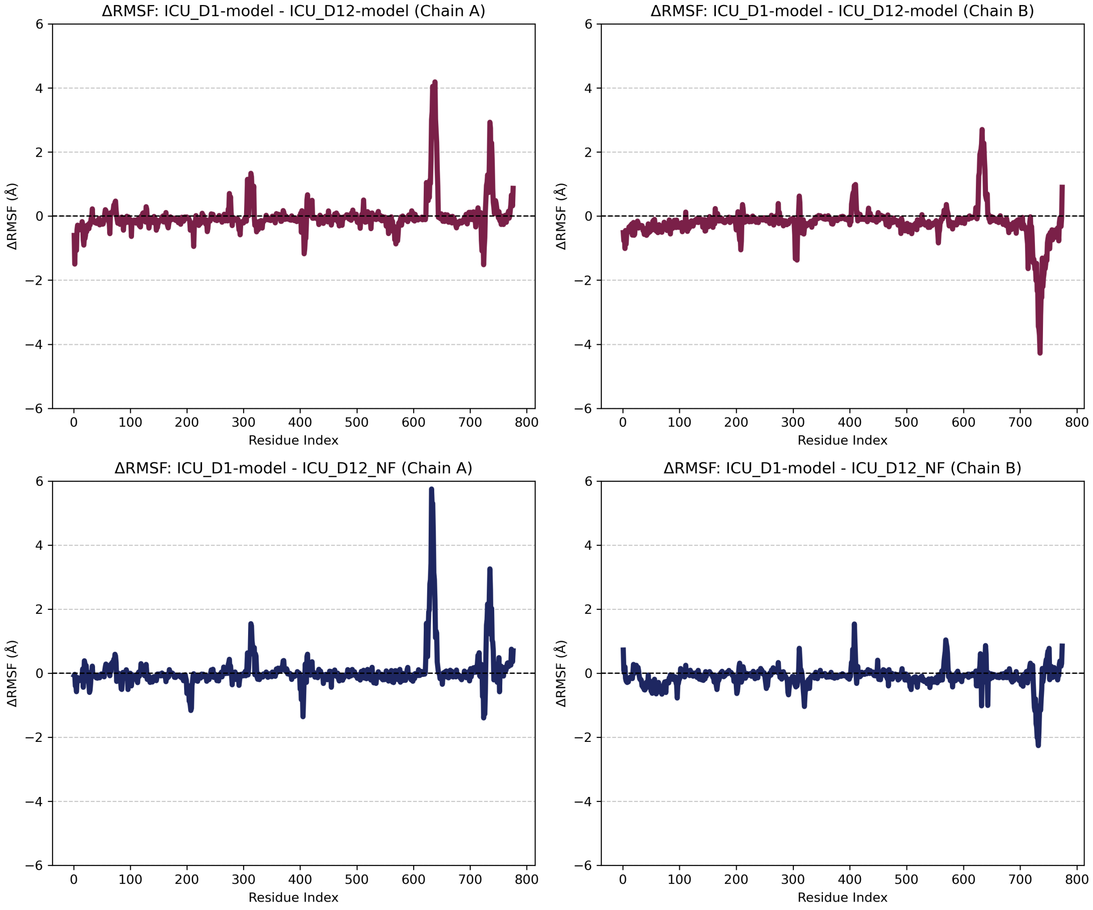
ΔRMSF: ICU_D1-model - ICU_D12-model (Chain A)
ΔRMSF: ICU_D1-model - ICU_D12-model (Chain B)
ΔRMSF: ICU_D1-model - ICU_D12_NF (Chain A)
ΔRMSF: ICU_D1-model - ICU_D12_NF (Chain B)
ΔRMSF (Å)
Residue Index
6
4
2
0
−2
−4
−6
0
100
200
300
400
500
600
700
800

**Figure E5.**

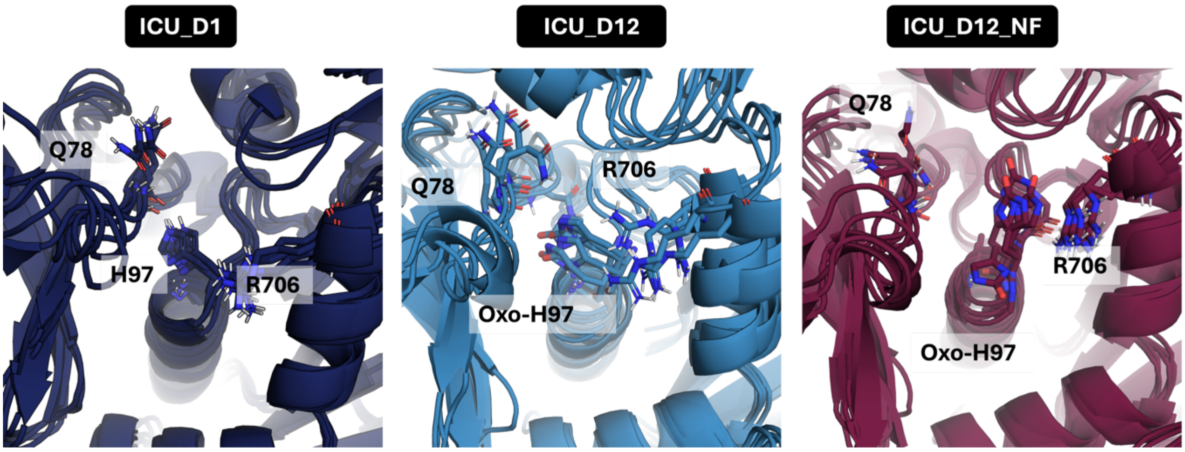
ICU_D1
ICU_D12
ICU_D12_NF
Q78
H97
R706
Q78
R706
Oxo-H97
Q78
R706
Oxo-H97

**Figure E6.**

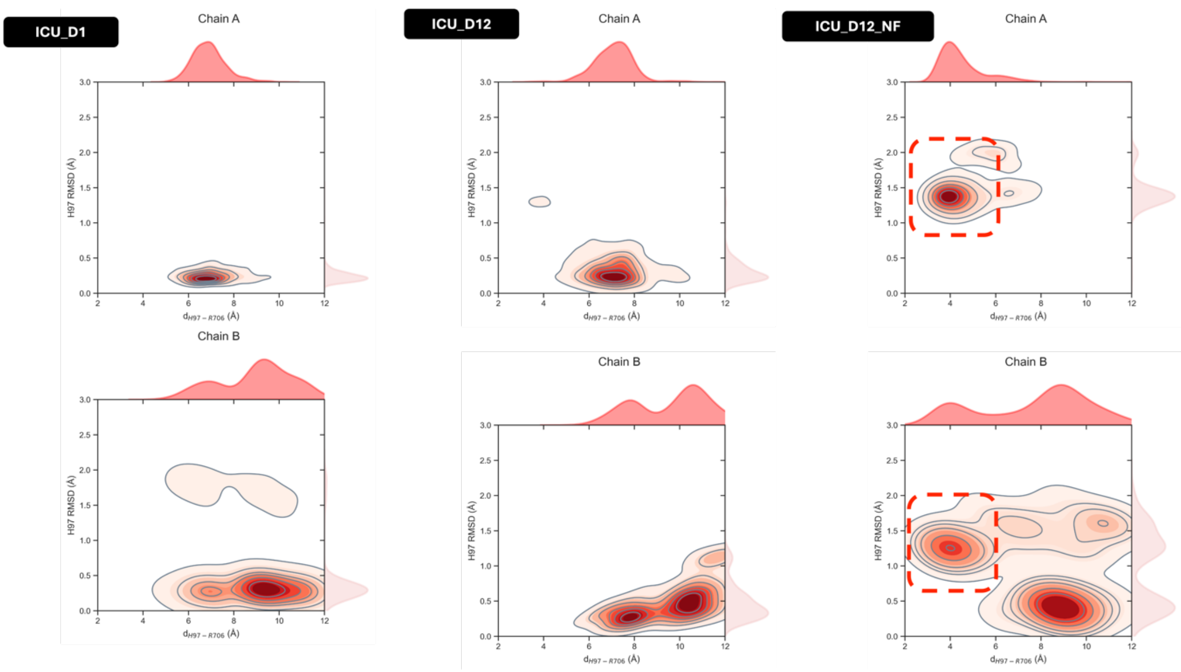

**Figure E7.**

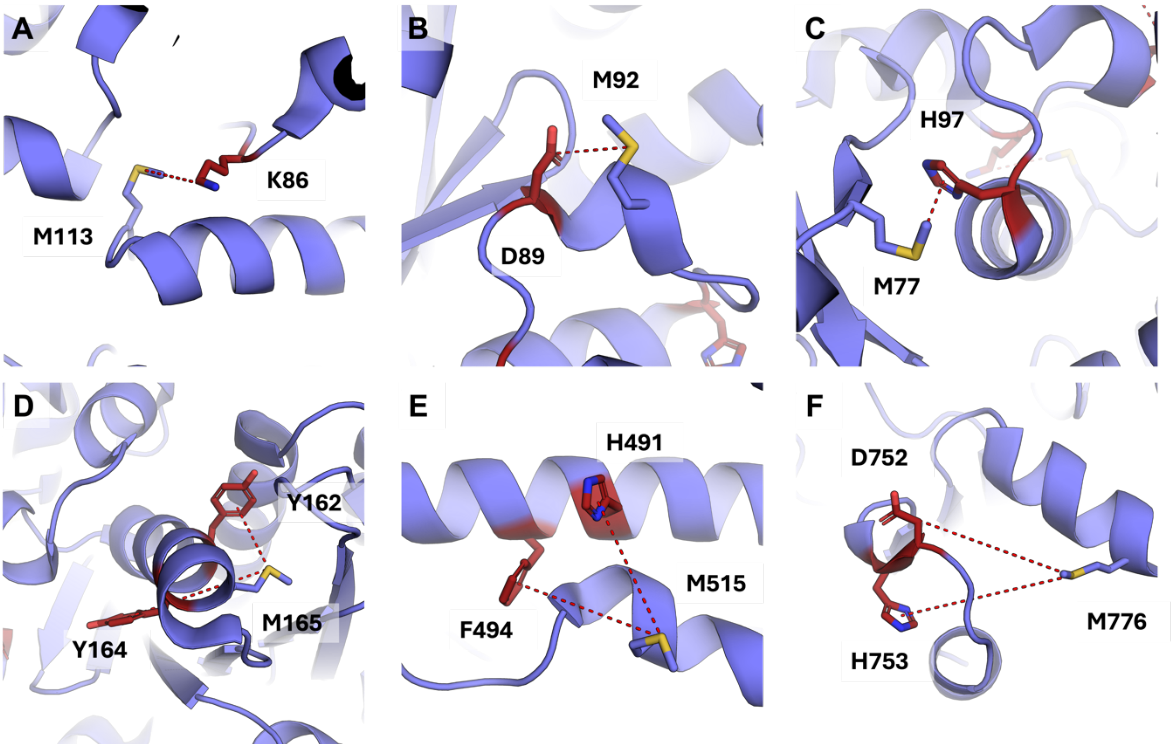
A
M113
K86
B
M92
D89
C
H97
M77
D
Y162
M165
Y164
E
H491
F494
M515
F
D752
M776
H753

**Figure E8.**

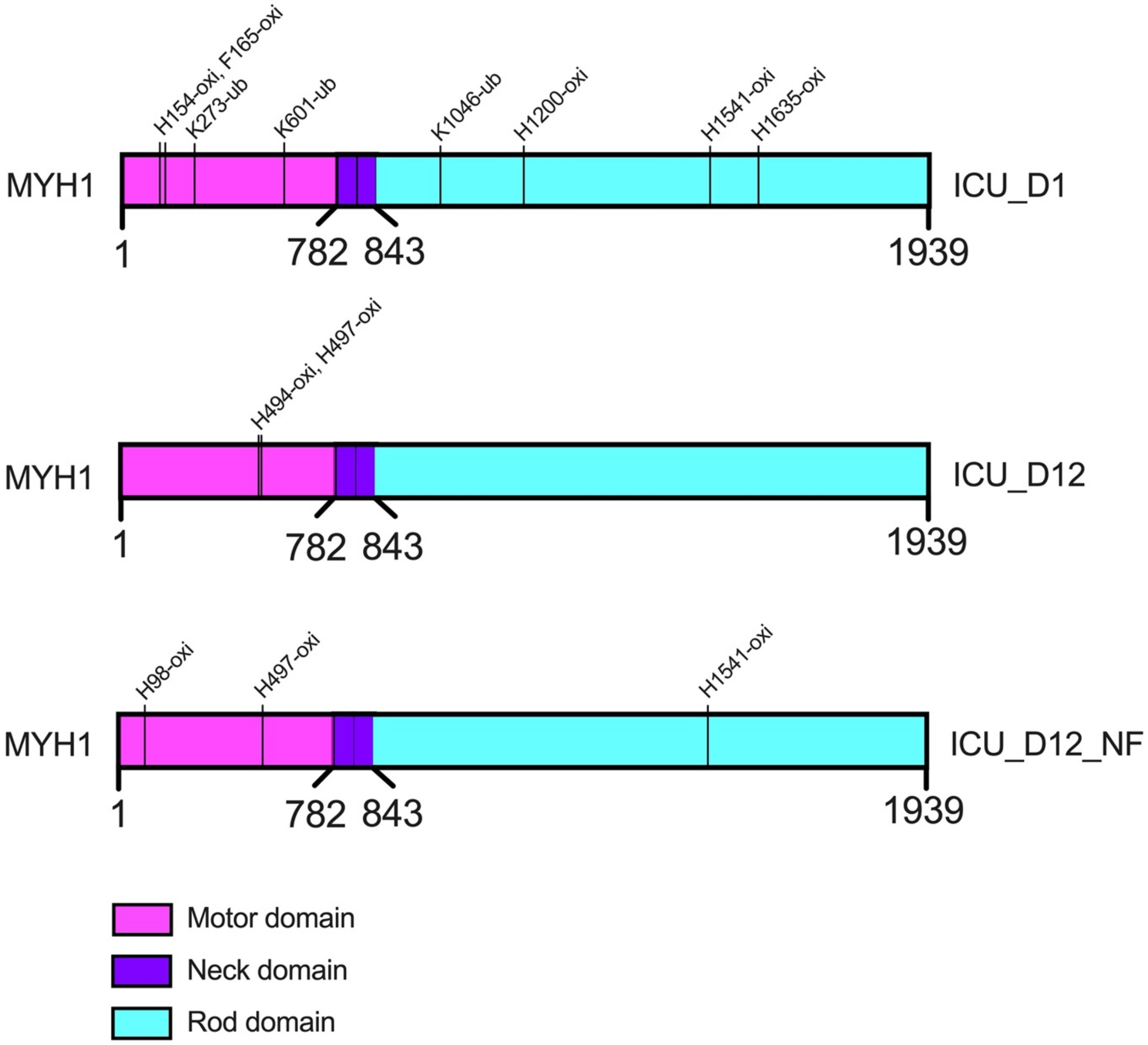
H154-oxi, F165-oxi
K273-ub
K601-ub
K1046-ub
H1200-oxi
H1541-oxi
H1635-oxi
MYH1
ICU_D1
1
782
843
1939
H494-oxi, H497-oxi
MYH1
ICU_D12
1
782
843
1939
H98-oxi
H497-oxi
H1541-oxi
MYH1
ICU_D12_NF
1
782
843
1939
Motor domain
Neck domain
Rod domain
